# Spatial and Temporal Drug Usage Patterns in Wastewater Correlate with Socioeconomic and Demographic Indicators in Southern Nevada

**DOI:** 10.1101/2024.02.02.24302241

**Authors:** Xiaowei Zhuang, Michael A. Moshi, Oscar Quinones, Rebecca A. Trenholm, Ching-Lan Chang, Dietmar Cordes, Brett J. Vanderford, Van Vo, Daniel Gerrity, Edwin C. Oh

**Author notes:** To whom correspondence should be addressed: Edwin Oh.

## Abstract

Evaluating drug use within populations in the United States poses significant challenges due to various social, ethical, and legal constraints, often impeding the collection of accurate and timely data. Here, we aimed to overcome these barriers by conducting a comprehensive analysis of drug consumption trends and measuring their association with socioeconomic and demographic factors. From May 2022 to April 2023, we analyzed 208 wastewater samples from eight sampling locations across six wastewater treatment plants in Southern Nevada, covering a population of 2.4 million residents with 50 million annual tourists. Using bi-weekly influent wastewater samples, we employed mass spectrometry to detect 39 analytes, including pharmaceuticals and personal care products (PPCPs) and high risk substances (HRS). Our results revealed a significant increase over time in the level of stimulants such as cocaine (pFDR=1.40×10^-10^) and opioids, particularly norfentanyl (pFDR =1.66×10^-12^), while PPCPs exhibited seasonal variation such as peak usage of DEET, an active ingredient in insect repellents, during the summer (pFDR =0.05). Wastewater from socioeconomically disadvantaged or rural areas, as determined by Area Deprivation Index (ADI) and Rural-Urban Commuting Area Codes (RUCA) scores, demonstrated distinct overall usage patterns, such as higher usage/concentration of HRS, including cocaine (p=0.05) and norfentanyl (p=1.64×10^-5^). Our approach offers a near real-time, comprehensive tool to assess drug consumption and personal care product usage at a community level, linking wastewater patterns to socioeconomic and demographic factors. This approach has the potential to significantly enhance public health monitoring strategies in the United States.

## Introduction

Monitoring drug consumption behaviors in the United States presents a complex challenge, both at individual and community levels^1,2^. Individuals often hesitate to self-report due to a variety of concerns encompassing social stigma, ethical dilemmas, privacy issues, and legal ramifications^3^. This reluctance can lead to biases that diminish the accuracy and reliability of collected data. At the community level, drug consumption behaviors are subject to rapid changes, often influenced by the emergence of new substances and the prevalence of polydrug use^4,5^. Furthermore, neighborhood characteristics—such as the degree of urbanization, demographic profiles, and social determinants of health—can significantly alter drug consumption patterns^6^. Historically, neighborhood disparities have been linked to various health-related behaviors, outcomes, and mortality^7–9^. Yet, the specific impact of urbanization and social determinants on drug consumption patterns remains an underexplored area. A deeper understanding of the interplay between drug consumption behaviors and socioeconomic factors could aid in identifying risk factors for drug overdoses and support efforts to promote health equity.

In response to COVID-19, wastewater monitoring programs have gained renewed importance as a method for tracking public health threats, providing real-time insights through the analysis of community sewage^10–12^. This method can detect a wide range of substances, from pharmaceuticals^2,13,14^ to pathogens^15–21^, thereby reflecting the health behaviors and exposures of a population. Such analysis reveals important trends in drug usage, dietary habits, and the presence of environmental contaminants—key indicators of social determinants like economic status, healthcare access, and environmental risks. Moreover, it can uncover health disparities across neighborhoods by examining substance concentrations that correlate with socioeconomic and lifestyle factors. For example, studies outside the United States have shown a link between socioeconomic or demographic factors and the consumption of specific chemicals or dietary components^6^. In the United States, tools like the Area Deprivation Index (ADI)^22,23^ and Rural-Urban Commuting Area (RUCA) codes^24^ provide in-depth insights into these factors. When used in conjunction with wastewater analytics, these tools have the potential to enable a detailed understanding of health disparities across neighborhoods.

In this study, we analyzed wastewater data from Southern Nevada over a span of 12 months to characterize drug consumption behaviors across a population of ∼2.4 million people and the ∼50 million tourists that visit Las Vegas annually. Using wastewater data on high risk substances (HRS) and pharmaceuticals and personal care products (PPCPs), we asked several questions: 1) Do drug consumption patterns cluster based on geographic locations, 2) Do consumption patterns change over time, and 3) Can socioeconomic variables be associated with the consumption of HRS and PPCPs. Taken together, our data highlight how wastewater data can be used to complement conventional public health tools and be leveraged for the analysis of population health dynamics in Southern Nevada.

## Results

### Spatial trends in drug usage patterns across Southern Nevada

To investigate spatial trends in drug usage and consumption, we conducted an unsupervised clustering analysis of analytes across various sewersheds (**Figure 1A**). For each facility, normalized usage rates of 34 metabolites (five metabolites were detected in less than 10% samples and therefore not included) across 26 time points were studied, resulting in a total of 884 measurements. Our results demonstrated that pairwise Pearson’s correlations (r) of drug usage among these facilities consistently exceeded 0.90, indicating significant similarities in usage trends (**Figure 1B**). The highest similarity was found between Facilities 1 and 3 (r=0.96), both serving larger populations, and between Facilities 4A and 4B (r=0.96), located within the same geographic community. In contrast, Facilities 5 and 6 displayed distinct patterns, with average correlations being 0.83±0.08 and 0.72±0.03, respectively, when compared to other facilities (**Figure 1B**). Although the patterns for PPCPs mirrored the overall trends for PPCPs combined with HRS (**Figure 1C**), the HRS-focused analysis showed significant differences, especially in Facilities 2 and 6 compared to other wastewater treatment plants (WWTPs) (**Figure 1D**).

**Figure 1.**
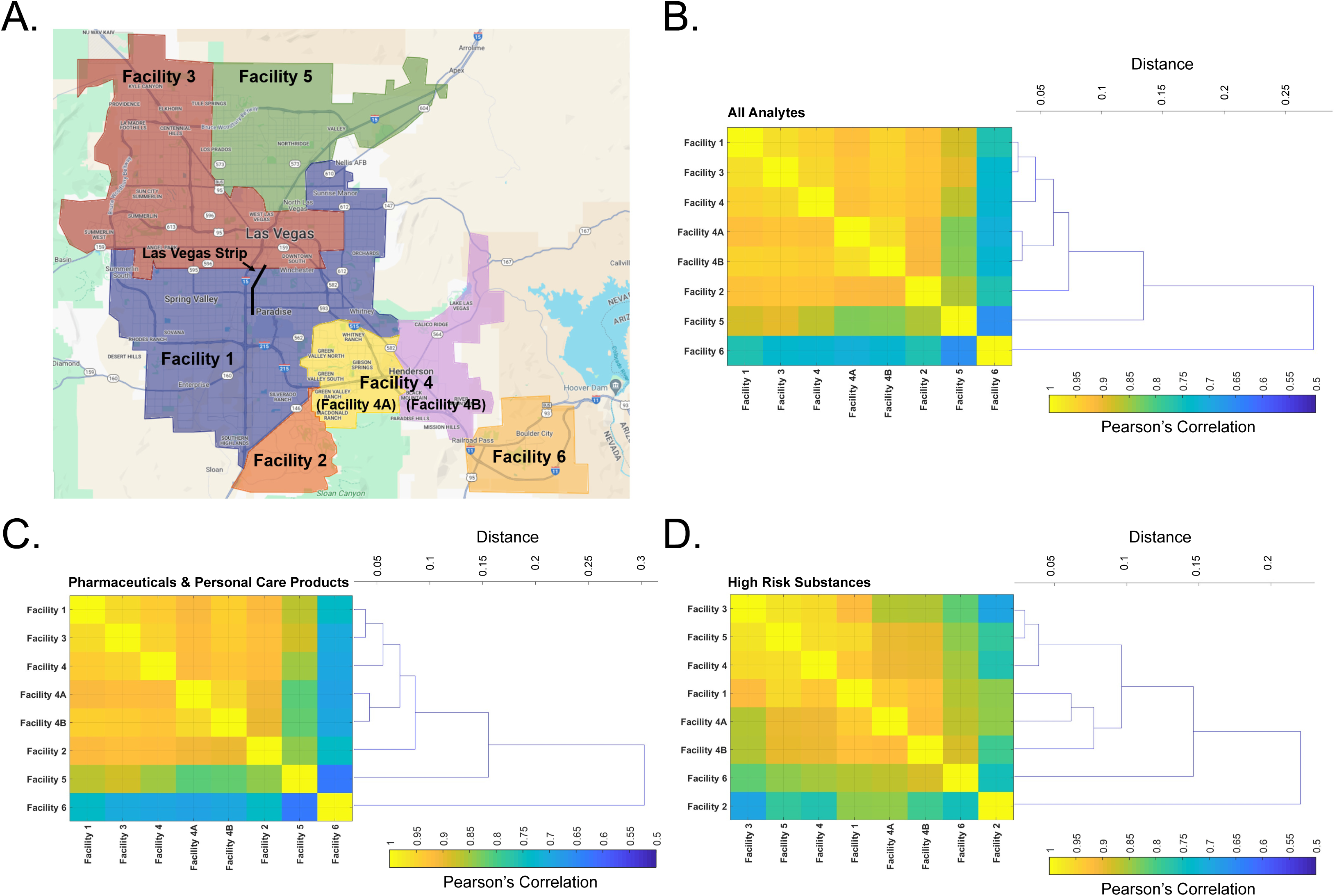
Spatial characteristics of pharmaceutical and personal care product (PPCP) and high risk substance (HRS) usage patterns across Southern Nevada sewersheds from May 2022 to Apr 2023. **(A)** Map of eight sampling locations across six wastewater treatment plants in Southern Nevada. **(B)** Similarities in usage patterns across eight locations, revealed by the Pearson’s correlation matrix for usage of all PPCPs and HRS (mg/day-person) from May 2022 to Apr 2023 (N_measure_=884 for each location). **(C)** Similarities in PPCP or **(D)** HRS usage patterns across facilities, revealed by the Pearson’s correlation matrix.

As a validation of our approach, we characterized the similarities for each analyte across 26 time points and 8 sampling locations. We found a robust correlation in the levels of cocaine and its metabolites, specifically ecgonine, ecgonine methyl ester, and benzoylecgonine (total of 208 measures for each analyte, r=0.87±0.09, first red box, **Figure 2A**). Pain relievers, including acetaminophen and the two nonsteroidal anti-inflammatory drugs (NSAIDs) (ibuprofen and naproxen), recreational marijuana metabolites (THC-COOH, THC-OH), and central nervous system (CNS) stimulants (amphetamine, methamphetamine) also showed closely related usage patterns (r=0.74±0.12, second red box). Opioids such as methadone (and its major metabolite 2-ethylidene-1,5-dimethyl-3,3-diphenylpyrrolidine (EDDP)), oxycodone, hydrocodone, and tramadol exhibited similar consumption trends (r=0.58±0.12, third red box), and correlated with the usage of the over-the-counter pain relievers. Additionally, our observations revealed interconnected usage patterns among specific PPCPs. This is highlighted by the significant correlation between caffeine and sucralose (r=0.67, fourth red box), as well as the frequent co-prescription of certain antibiotics, such as sulfamethoxazole and trimethoprim (r=0.69, fifth red box) (**Figure 2A**).

**Figure 2.**
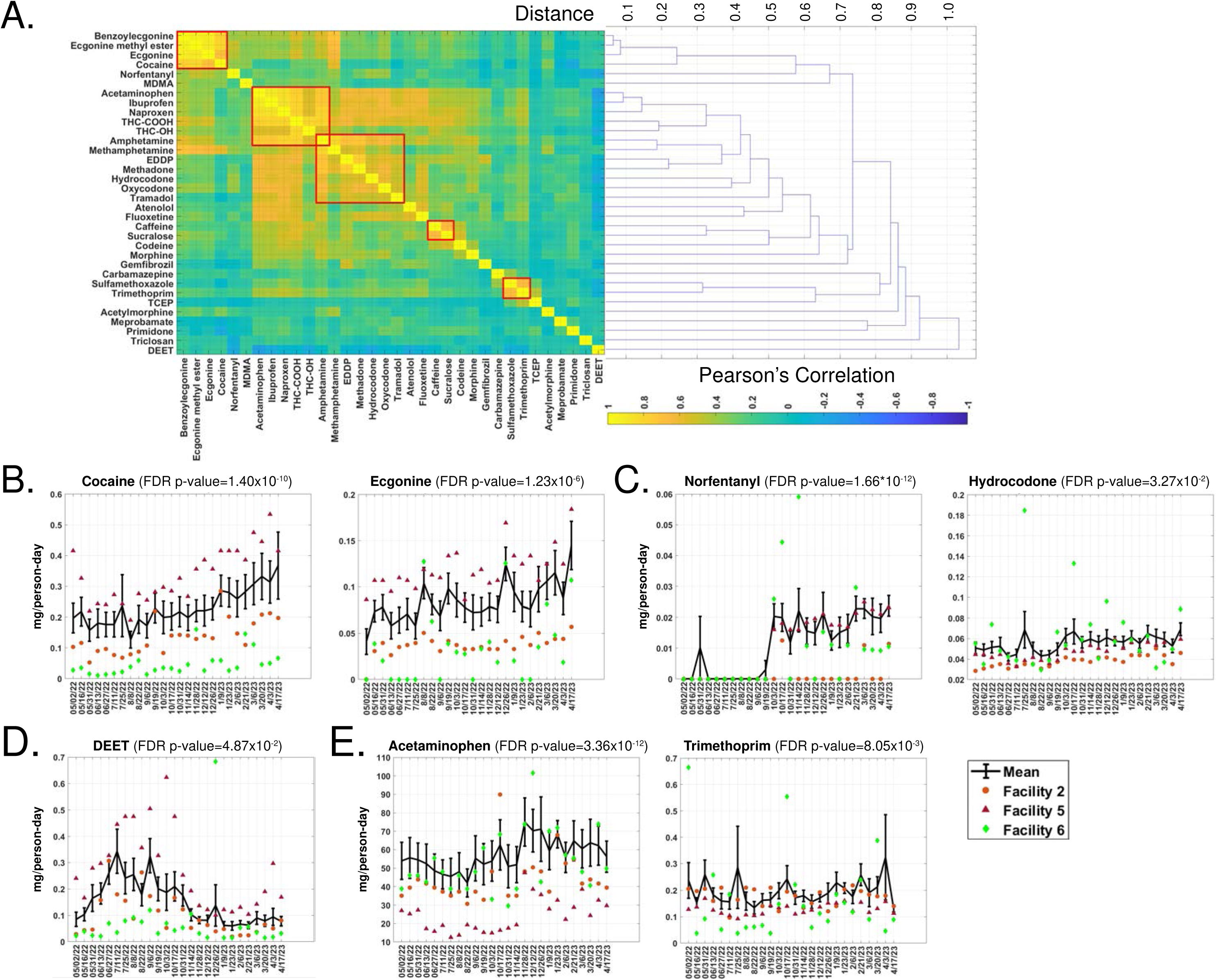
Temporal characteristics of usage patterns across all pharmaceuticals and personal care products (PPCPs) and high risk substances (HRS) in Southern Nevada sewersheds from May 2022 to Apr 2023. **(A)** Similarities in consumption (mg/day-person) patterns across all PPCPs and HRS, revealed by the Pearson’s correlation matrix for each analyte across eight sampling locations from May 2022 to Apr 2023 (N_measure_=208 for each analyte). Five red boxes, from upper left to bottom right, indicate similar usage/occurrence patterns of cocaine-related metabolites, pain relievers, opioids, daily PPCPs and prescribed antibiotics. **(B-E)** Significant temporal trends in the linear mixed model for seven analytes, including: **(B)** increased consumption of cocaine (and occurrence of its metabolite ecgonine) from May 2022 to 2023; **(C)** increased consumption of opioids, such as fentanyl (based on its major metabolite norfentanyl) and hydrocodone, from Sep. 2022 to 2023; and **(D-E)** PPCP usage patterns fluctuated significantly and revealed seasonal consumption patterns.

### Temporal trends in drug usage patterns

In our longitudinal study, with repeated measurements of PPCPs and HRS-related analytes, we sought to identify temporal trends in usage and consumption patterns. Utilizing a linear mixed effect (LME) model, we discovered a significant time-related effect for nine of the 34 analytes. This included six HRS (**Figure 2B-C** and **Supplementary** Figure 1) and three PPCPs (**Figure 2D-E**).

Among the HRS, cocaine occurrence exhibited a steady rise over time (*p_FDR_*=1.40×10^-10^), as did its major metabolites ecgonine, ecgonine methyl ester, and benzoylecgonine, indicating a marked increase from 2022 to 2023 (**Figure 2B** and **Supplementary** Figure 1). Prior to September 2022, Norfentanyl was detected only on Memorial Day weekend in May 2022 at Facility 4A, but a significant increase in detection frequency and concentration was observed at all facilities starting in September/October 2022 (*p_FDR_* =1.66×10^-12^, **Figure 2C**). A direct comparison for Facility 1 suggested an increase in occurrence/consumption between 2010^13^ and 2023 for 14 HRS (green in Supplementary Table 1).

Unlike for the HRS, the usage patterns of PPCPs showed significant temporal fluctuations between 2022 and 2023. *N,N*-Diethyl-meta-toluamide (DEET), an insect repellent ingredient, peaked in the summer of 2022 and declined towards spring 2023 (*p_FDR_*=4.87×10^-2^, **Figure 2D**), reflecting a seasonal usage pattern. Acetaminophen usage surged in November 2022, remaining high through the holiday season until January 2023 (*p_FDR_*=3.36×10^-2^, **Figure 2E**). The level of PPCP usage in 2023 paralleled those in 2010^13^ (yellow in **Supplementary Table 1**), including atenolol, primidone, carbamazepine, trimethoprim, sulfamethoxazole, and DEET. Interestingly, a decline was observed in the use of meprobamate, a popular sedative in the 1950s, and tris (2-chloroethyl) phosphate (TCEP), a flame retardant, compared to 2010 (red in **Supplementary Table 1**). Overall, our analysis highlights an uptick in HRS consumption and seasonal variation in PPCP use/occurrence in Southern Nevada.

### Correlating PPCP and HRS Usage Patterns with Neighborhood Context

Our retrospective analysis of the urban-rural status and neighborhood contexts of eight sampling locations revealed two key findings. First, Facility 6 was unique with a higher Rural-Urban Commuting Area (RUCA) code of 2, indicating less urbanization compared to other facilities (**Table 1**). Second, Facility 5 had a significantly higher Area Deprivation Index (ADI) than Facility 2, suggesting a more socioeconomically disadvantaged population (**Figure 3**). The LME model showed a significant location or location x time effect for nearly all analytes, except for the antibiotics trimethoprim and sulfamethoxazole, TCEP, and triclosan (**Table 2**), highlighting distinct spatial trends across the facilities. The significant post-hoc pairwise differences among Facilities 2, 5 and 6 further linked the distinct analyte occurrence patterns to neighborhood contexts (**Table 3**). For HRS, significant consumption pattern differences were observed between facilities, with Facilities 2 and 5 typically showing the lowest and highest rates, respectively. In contrast, fewer PPCPs showed significant differences between facilities (**Table 3**).

**Figure 3.**
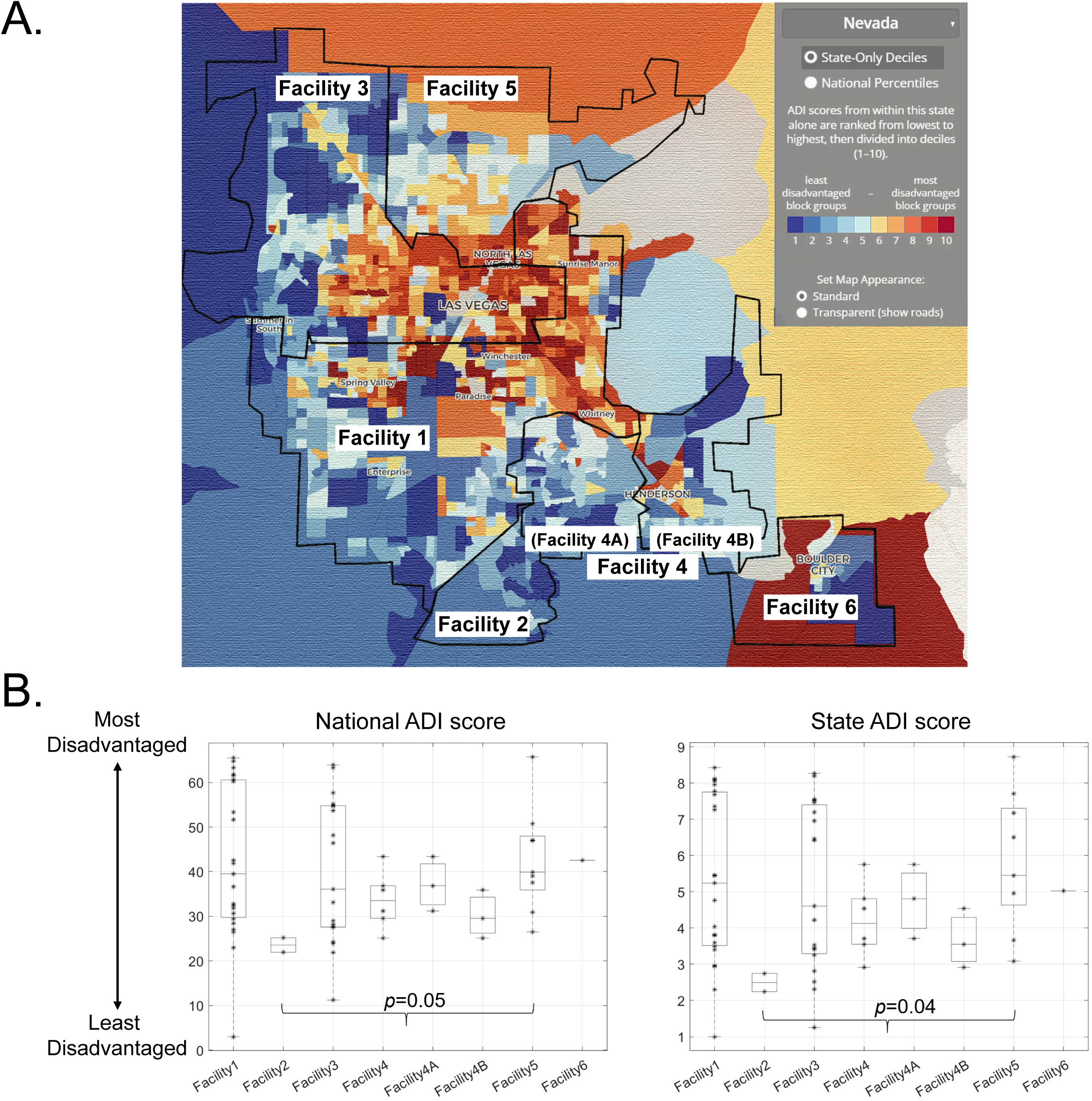
Correlating rankings of neighborhoods by socioeconomic disadvantage with wastewater facilities. **(A)** Comparisons of neighborhood context, in terms of the national and state area deprivation index (ADI), across each **(B)** Southern Nevada sewershed.

**Table 1.** Southern Nevada sewershed coverages by zip codes. Average daily flow (million gallons per day (mgd)) and sewershed population (number of people) are listed below each facility. Socioeconomic and demographic characteristics of each sewershed are characterized using 2010 rural-urban commuting area codes (RUCA) and area deprivation index (ADI) scores, respectively. A RUCA of 1 represents metropolitan area core: primary flow within an urbanized area (UA), and a RUCA of 2 represents metropolitan area high commuting: primary flow 30% or more to a UA. The ADI allows for rankings of neighborhood by socioeconomic disadvantage at the national (National ADI) and state (State ADI) levels. A higher ADI score indicates more socioeconomic disadvantages. ADI is based on 9-digit zip codes, and therefore an average value is computed and listed for each 5-digit zip code here.

|  | Zip-code | Ruca1 | National ADI | State ADI |  | Zip-code | Ruca1 | National ADI | State ADI |
| --- | --- | --- | --- | --- | --- | --- | --- | --- | --- |
| Facility 1 | 89103 | 1 | 61.51 | 7.27 | Facility 2 | 89052 | 1 | 25.17 | 2.75 |
| 100 mgd | 89109 | 1 | 32.78 | 4.03 | 5 mgd | 89044 | 1 | 21.93 | 2.24 |
| 872,009 | 89113 | 1 | 29.43 | 3.49 | 86,330 |  |  |  |  |
|  | 89115 | 1 | 64.74 | 8.10 | Facility 3 | 89101 | 1 | 63.90 | 8.19 |
|  | 89118 | 1 | 31.82 | 3.81 | 42 mgd | 89104 | 1 | 54.81 | 7.20 |
|  | 89119 | 1 | 61.81 | 7.96 | 757,418 | 89110 | 1 | 57.66 | 7.55 |
|  | 89120 | 1 | 42.51 | 5.45 |  | 89143 | 1 | 28.95 | 3.45 |
|  | 89121 | 1 | 63.31 | 8.05 |  | 89131 | 1 | 24.28 | 2.81 |
|  | 89122 | 1 | 65.50 | 8.43 |  | 89166 | 1 | 23.92 | 2.31 |
|  | 89123 | 1 | 32.35 | 3.81 |  | 89149 | 1 | 27.53 | 3.42 |
|  | 89139 | 1 | 27.03 | 2.95 |  | 89130 | 1 | 36.06 | 4.61 |
|  | 89141 | 1 | 22.94 | 2.30 |  | 89129 | 1 | 33.08 | 4.23 |
|  | 89142 | 1 | 51.69 | 7.36 |  | 89138 | 1 | 11.22 | 1.25 |
|  | 89146 | 1 | 41.89 | 5.46 |  | 89134 | 1 | 28.27 | 3.53 |
|  | 89147 | 1 | 39.53 | 5.24 |  | 89144 | 1 | 21.87 | 2.52 |
|  | 89148 | 1 | 30.67 | 3.60 |  | 89145 | 1 | 46.44 | 6.43 |
|  | 89156 | 1 | 60.64 | 8.12 |  | 89128 | 1 | 48.14 | 6.46 |
|  | 89158 | 1 | 3.00 | 1.00 |  | 89108 | 1 | 53.69 | 7.53 |
|  | 89169 | 1 | 60.29 | 7.78 |  | 89107 | 1 | 54.71 | 7.47 |
|  | 89178 | 1 | 26.50 | 2.96 |  | 89102 | 1 | 55.16 | 6.95 |
|  | 89179 | 1 | 28.44 | 3.40 |  | 89106 | 1 | 63.25 | 8.27 |
|  | 89183 | 1 | 36.66 | 4.77 |  | 89117 | 1 | 27.73 | 3.25 |
|  | 89199 | 1 | 53.30 | 7.69 | Facility 4A | 89011 | 1 | 36.83 | 4.81 |
| Facility 5 | 89085 | 1 | 26.49 | 3.09 | 16 mgd | 89015 | 1 | 43.39 | 5.75 |
| 20 mgd | 89084 | 1 | 30.91 | 3.66 | 133,977 | 89002 | 1 | 31.18 | 3.72 |
| 255,008 | 89031 | 1 | 39.90 | 5.45 | Facility 4B | 89014 | 1 | 35.86 | 4.54 |
|  | 89032 | 1 | 46.95 | 6.50 | 6 mgd | 89074 | 1 | 29.54 | 3.55 |
|  | 89030 | 1 | 65.73 | 8.73 | 114,532 | 89012 | 1 | 25.12 | 2.92 |
|  | 89081 | 1 | 37.57 | 4.96 |  |  |  |  |  |
|  | 89086 | 1 | 39.04 | 4.96 | Facility 6 |  |  |  |  |
|  | 89033 | 1 | 47.04 | 7.17 | 0.8 mgd | 89005 | 2 | 42.55 | 5.03 |
|  | 89165 | 1 | 50.71 | 7.71 | 16,399 |  |  |  |  |

**Table 2.** F-Statistics and significance levels (*p*-values) of time, facility, and time-facility interactions in the linear mixed effect model running using data from all eight facilities. P values are corrected for multiple comparisons using the false-discovery rate (FDR) method. Degrees of freedom (dF) are listed below each F-statistics.

| Name | PPCP/HRS | Category | Fvalue: Facility<br>dF(7,72) | FDR-p: Facility | Fvalue: Time<br>dF(1,72) | FDR-p:<br>Time | Fvalue:<br>Interaction<br>dF(7,72) | FDR-p:<br>Interaction |
| --- | --- | --- | --- | --- | --- | --- | --- | --- |
| Acetaminophen | PPCP | OTC Pain Reliever | 21.63 | 3.70E-20 | 6.13 | 3.36E-02 |  |  |
| Ibuprofen | PPCP | NSAID | 20.84 | 1.44E-19 |  |  |  |  |
| Naproxen | PPCP | NSAID | 24.63 | 1.76E-22 |  |  |  |  |
| Atenolol | PPCP | Licit Drug | 14.34 | 5.10E-14 |  |  |  |  |
| Carbamazepine | PPCP | Licit Drug | 5.91 | 1.31E-05 |  |  | 3.22 | 8.79E-03 |
| Fluoxetine | PPCP | Licit Drug | 10.04 | 6.36E-10 |  |  |  |  |
| Gemfibrozil | PPCP | Licit Drug | 2.72 | 2.64E-02 |  |  |  |  |
| Meprobamate | PPCP | Licit Drug | 3.04 | 1.25E-02 |  |  |  |  |
| Primidone | PPCP | Licit Drug | 2.63 | 3.18E-02 |  |  |  |  |
| Sulfamethoxazole | PPCP | Licit Drug |  |  |  |  |  |  |
| Trimethoprim | PPCP | Licit Drug |  |  | 9.31 | 8.05E-03 |  |  |
| Caffeine | PPCP | Daily | 3.63 | 3.57E-03 |  |  |  |  |
| DEET | PPCP | Daily | 6.62 | 2.28E-06 | 5.37 | 4.87E-02 |  |  |
| Sucralose | PPCP | Daily | 3.19 | 8.89E-03 |  |  |  |  |
| TCEP | PPCP | Daily |  |  |  |  |  |  |
| Triclosan | PPCP | Daily |  |  |  |  |  |  |
| Acetylmorphine | HRS | Opioid | 3.24 | 8.52E-03 |  |  |  |  |
| Codeine | HRS | Opioid | 2.87 | 1.89E-02 |  |  |  |  |
| EDDP | HRS | Opioid | 14.22 | 6.21E-14 |  |  |  |  |
| Hydrocodone | HRS | Opioid | 8.78 | 1.14E-08 | 6.22 | 3.27E-02 |  |  |
| MDMA | HRS | Opioid | 5.83 | 1.55E-05 |  |  |  |  |
| Methadone | HRS | Opioid | 26.03 | 1.86E-23 |  |  |  |  |
| Morphine | HRS | Opioid | 3.77 | 2.61E-03 |  |  | 3.19 | 8.89E-03 |
| Norfentanyl | HRS | Opioid |  |  | 62.20 | 1.66E-12 | 4.52 | 4.16E-04 |
| Oxycodone | HRS | Opioid | 15.53 | 4.72E-15 |  |  |  |  |
| Tramadol | HRS | Opioid | 17.22 | 1.68E-16 |  |  |  |  |
| Amphetamine | HRS | Stimulant | 48.84 | 2.21E-37 |  |  | 2.51 | 4.03E-02 |
| Benzoyllecgonine | HRS | Stimulant | 16.85 | 3.22E-16 | 14.62 | 6.49E-04 |  |  |
| Cocaine | HRS | Stimulant | 14.84 | 1.88E-14 | 50.59 | 1.40E-10 | 3.29 | 7.99E-03 |
| Ecgonine | HRS | Stimulant | 10.00 | 6.66E-10 | 28.48 | 1.23E-06 | 3.57 | 4.06E-03 |
| Ecgonine methyl ester | HRS | Stimulant | 10.60 | 1.77E-10 | 51.09 | 1.22E-10 | 4.78 | 2.23E-04 |
| Methamphetamine | HRS | Stimulant | 45.26 | 1.05E-35 |  |  |  |  |
| THC-COOH | HRS | Marijuana | 28.34 | 4.72E-25 |  |  |  |  |
| THC-OH | HRS | Marijuana | 9.06 | 6.12E-09 |  |  |  |  |

**Table 3.**
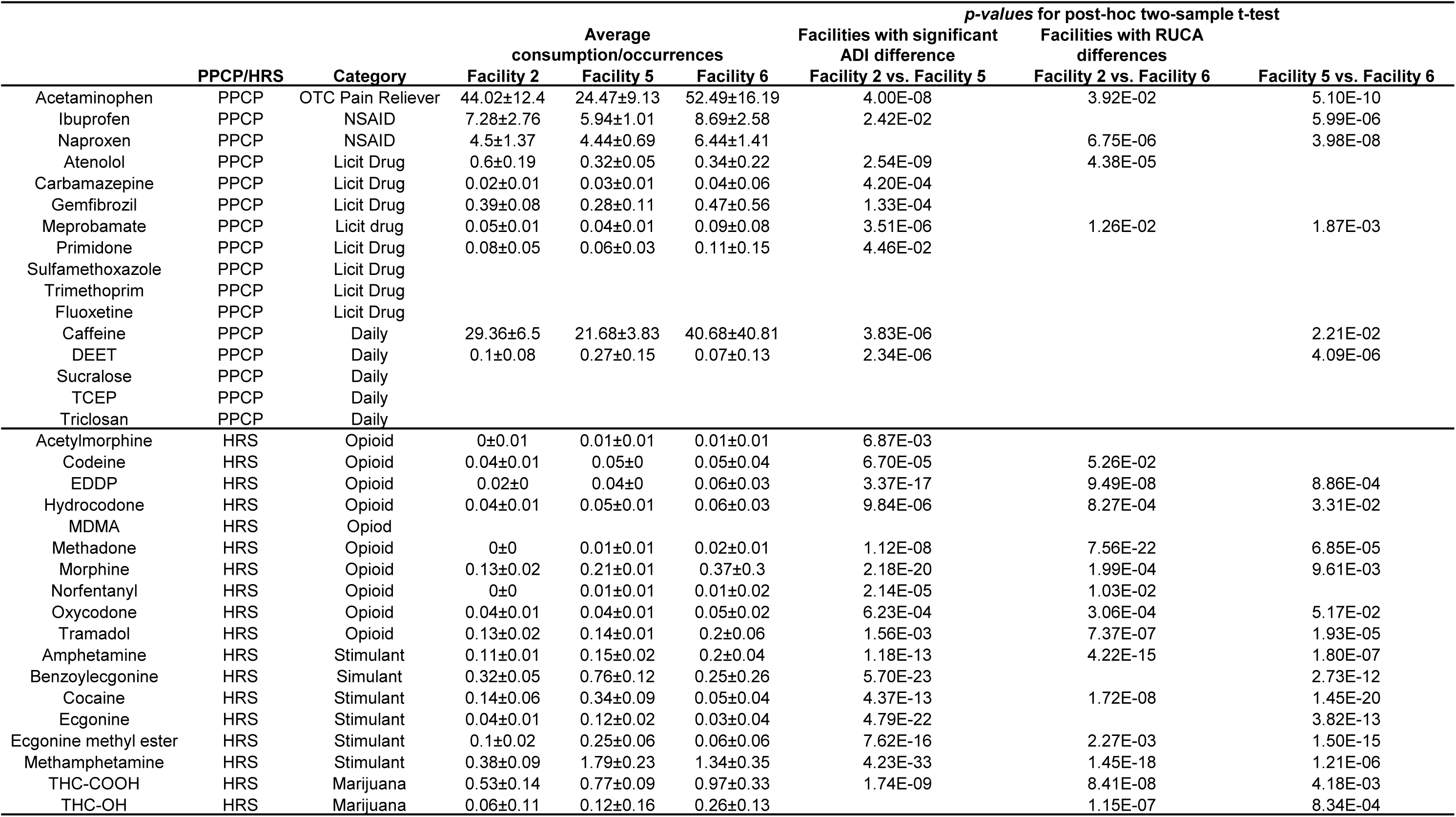
Post-hoc comparisons among Facilities 2, 5 and 6 that share significantly different RUCA code and neighborhood contexts. Average consumptions across time are listed in column 4 to 6, and significant levels (*p*-values) for pair-wise between facility comparisons are listed in column 7 to 9.

Among the 30 analytes that displayed a significant effect based on location or location x time (**Supplementary** Figure 2-4), the consumption/occurrence patterns of six analytes exhibited a significant positive correlation with the average ADI of each facility (i.e., more disadvantaged). This included cocaine and its metabolites ecgonine and benzoylecgonine (**Figure 4A**), as well as methamphetamine, norfentanyl (major metabolite of fentanyl), and the anti-convulsant carbamazepine (**Figure 4B**). These associations further underscore the relationship between drug usage patterns in different facilities and socioeconomic factors.

**Figure 4.**
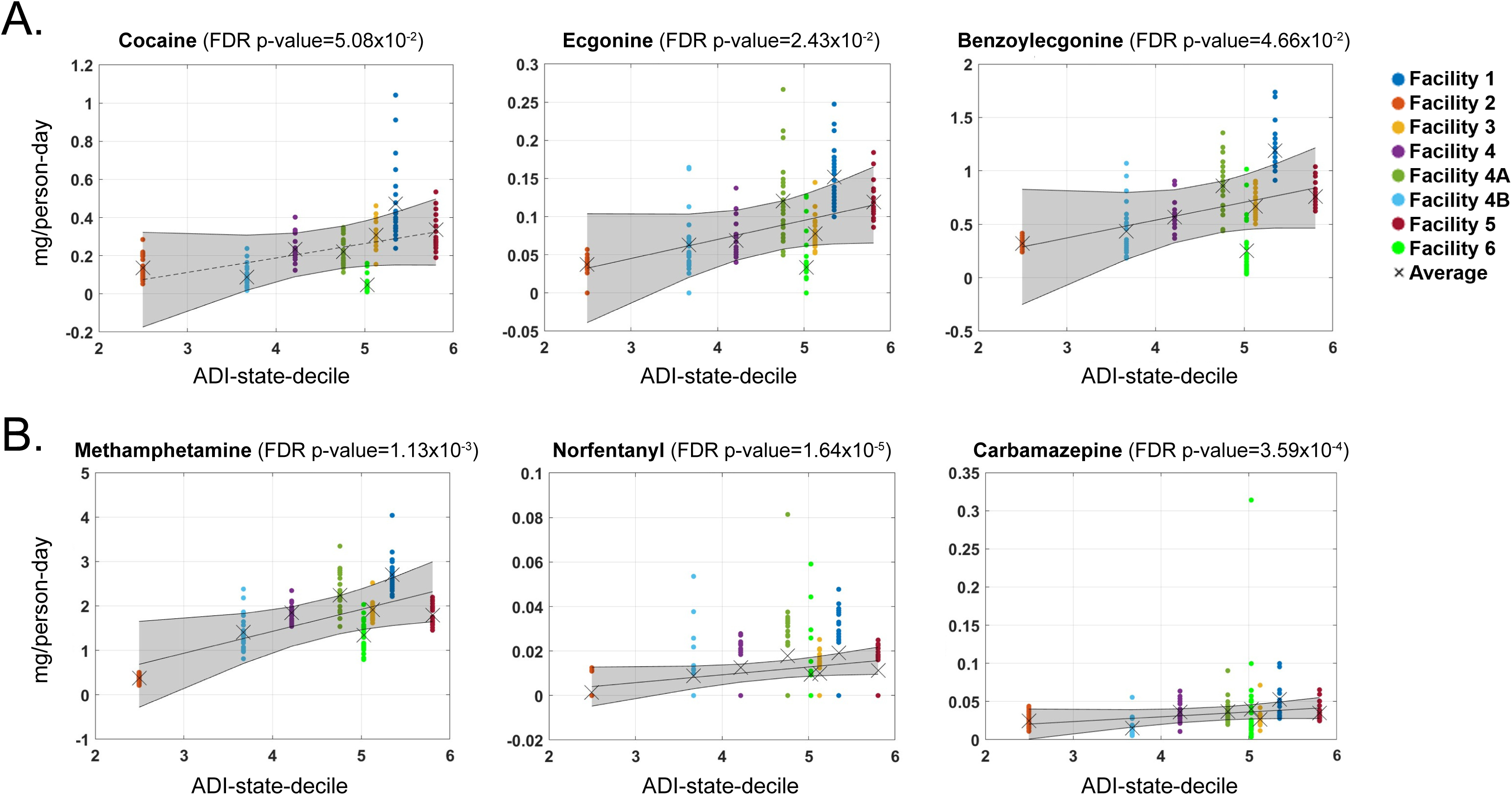
Significant association between drug usage (in mg/day-person) and neighborhood context revealed by area deprivation index (ADI). An increased usage with advanced neighborhood disadvantage were evident for **(A)** cocaine and its metabolites and **(B)** methamphetamine, norfentanyl, and carbamazepine. Circles (O) represent consumption values at each individual time point and crosses (X) indicate the average usage over time.

## Discussion

Wastewater monitoring is emerging as an innovative tool to address the growing problem of drug abuse^6,14,25^. In our study, we investigated the temporal and spatial patterns of 16 PPCPs and 18 HRS across eight sampling locations in Southern Nevada from May 2022 to April 2023. Our analysis revealed significant temporal variations in the estimated drug consumption from wastewater, highlighting an overall increase in HRS usage over time, alongside seasonally fluctuating PPCP utilization patterns. Moreover, by correlating wastewater drug consumption data with neighborhood contexts, we observed significantly greater HRS usage in more disadvantaged areas, as determined by ADI. These findings underscore the potential for wastewater monitoring programs to not only serve as a reliable method for tracking drug consumption, but also as a tool for identifying specific drug use patterns influenced by the socioeconomic and demographic characteristics of communities.

The unsupervised clustering analysis of drugs in wastewater revealed strong correlations in estimated consumption patterns, specifically among cocaine and its metabolites, between methadone and its derivative, and between methamphetamine and its partially excreted form, amphetamine (**Figure 2A**). These findings support the utility of wastewater data as a complementary source of information to determine drug exposure. Moreover, the clustering analysis of facilities highlighted geographically varied usage patterns, aligning with s socioeconomic and demographic differences across facilities, even without prior information on the neighborhoods they serve (**Figure 1B** and **Figure 3**). These findings imply that drug consumption patterns in wastewater at individual WWTPs are influenced by distinct community characteristics, laying a foundation for further exploration of how social determinants of health correlate with drug usage patterns in wastewater.

Our study revealed a significant increase in all four cocaine-related analytes across eight sampling locations over the past year (**Figure 2B**), including a substantial 70% increase in their usage at Facility 1 over the last decade (**Supplementary Table 1**). These wastewater data align with the increase in cocaine-related emergency department visits and hospital admissions reported in the 2022 Nevada Epidemiologic Profile^26^, underscoring the effectiveness and reliability of wastewater data in tracking drug consumption trends. Furthermore, consistent with previous studies linking cocaine use to income status in different geographic areas^6,27,28^, our application of the state-ranked ADI to assess regional socioeconomic conditions (encompassing education, income, housing, and household characteristics) also supports these findings. Spatial analysis revealed that communities with lower socioeconomic standing tend to show higher wastewater-estimated cocaine usage (**Figure 4A**), a pattern similarly observed in the increased use of another central nervous system (CNS) stimulant, methamphetamine, in these more disadvantaged neighborhoods (**Figure 4B**).

Wastewater-estimated opioid usage was strongly correlated with the consumption of NSAIDs, marijuana, and CNS stimulants (**Figure 2A**), suggesting facility-level polydrug usage often linked to chronic pain management. While opioid usage increased significantly in 2023 compared to 2010 (**Supplementary Table 1**), only norfentanyl and hydrocodone showed temporal changes from 2022 to 2023 (**Figure 2C**), indicating that a one-year interval might be insufficient to observe substantial variations in opioid usage. This could also be attributed to the diversity of available drugs influencing the fluctuation in individual opioid consumption. A detailed analysis of norfentanyl trends, reported in Gerrity et al., 2024, showed a significant increase post-October 2022, consistent with local clinical reports of fentanyl-related deaths in Southern Nevada^14^. Similar to cocaine, increased norfentanyl usage was significantly associated with neighborhoods facing socioeconomic disadvantages (**Figure 4B**), aligning with previous findings that higher prescription opioid rates correlate with social determinants of health such as poverty, unemployment, lower education levels, and unstable housing, both in the United States and internationally^6,27,28^. Given these findings and the observed increase in HRS in disadvantaged neighborhoods, combined with lifestyle challenges and limited healthcare resources in these areas^29^, wastewater monitoring of HRS could inform long-term public health planning in these communities.

Our clustering analysis of WWTPs using HRS revealed that Facility 2, characterized by a higher-income demographic and a large retirement-age population^14^, was the most distinct, followed by Facility 6 (**Figure 1D**). Consistent with having the lowest ADI in this study, Facility 2 exhibited the lowest consumption rates for nearly every HRS (**Figure 4**, **Table 3**, and **Supplementary** Figure 3), reinforcing the link between HRS usage and neighborhood socioeconomic status. A closer look at Facility 6 showed lower consumption of CNS stimulants and moderate use of opioids and marijuana **(Supplementary** Figure 3). Unique in its RUCA code of 2, these patterns in Facility 6 suggest that drug usage is influenced not only by socioeconomic factors but also by socio-demographic elements like urbanization.

Significant temporal changes were recorded in only three of the 16 PPCPs analyzed. In contrast to HRS, most PPCP usage remained stable when compared to 2010 data, indicating a generally consistent consumption over time **(Supplementary Table 1)**. The use of DEET, an insect repellent, exhibited marked seasonal variation with a peak in summer months (**Figure 2D**), affirming the reliability of wastewater data for monitoring PPCP usage. Additionally, the consumption of the acetaminophen varied quarterly, with an increase during the holiday season, while antibiotics like trimethoprim showed significant bi-monthly fluctuations (**Figure 2E**), suggesting regular and periodic use of these substances. Across different catchment areas, we observed significant variations in PPCP consumption/occurrence for all analytes except antibiotics (sulfamethoxazole and trimethoprim), TCEP (a flame retardant), and triclosan (an antimicrobial agent found in some soaps and lotions, **Table 2**). Interestingly, only the use of the anticonvulsant carbamazepine, like HRS, showed a significant positive correlation with the ADI (**Figure 4B**), possibly due to its use in treating neuropathic pain^6^. These patterns imply a relatively uniform use of PPCPs across socioeconomic strata, or alternatively, suggest that factors other than socioeconomic status, as indicated by ADI, may influence PPCP usage at specific facilities.

## Limitations

This study faces several limitations. First, the ADI offers refined resolution at 9-digit zip code levels, but our wastewater facilities cover broader areas spanning multiple 5-digit zip codes, leading to a generalized rather than precise socioeconomic status estimation for each facility. Although the RUCA codes are specific to 5-digit zip codes, uniform RUCA codes across all facilities simplified urbanization characterization. Notably, Facilities 6 and 2, serving fewer zip code areas, provided a more reliable SES context using ADI, reflected in the distinct consumption patterns of HRS observed in these facilities. Second, our analysis assumes that drug concentrations in wastewater solely represent population consumption, a premise challenged by factors like method sensitivity, in-sewer transformation, and alternative drug disposal methods. Third, our LME model only captures linear relationships between drug use and ADI scores or temporal changes, suggesting that more advanced multivariate and nonlinear methods could better assess complex associations. Finally, Facility 1 serves the Las Vegas Strip, an area known for tourism attractions and hospitality. Due to the mixing of analytes from tourists and the local population, our current analysis of Facility 1 is likely influenced by the confounding effects of mobile populations.

## Conclusions

To our knowledge, this is the first report to examine how spatiotemporal drug usage behaviors, examined through community wastewater, can be integrated with ADI or RUCA scores in the United States. The results of this observational study demonstrate how wastewater data can complement public health tools to provide an unbiased estimate of socioeconomic and demographic indicators in communities served by a wastewater treatment plant.

## Methods

### Data source

This observational study adhered to the Strengthening the Reporting of Observational Studies in Epidemiology (STROBE) reporting guidelines^30^. The University of Nevada Las Vegas (UNLV) Institutional Review Board (IRB) reviewed this project and determined it to be exempt from human subject research according to federal regulations and University policy.

### Wastewater collection and analysis

For this study, the methodology for wastewater collection and analysis by liquid chromatography tandem mass spectrometry (LC-MS/MS) with isotope dilution was extensively detailed in Gerrity et al., 2024^14^. Briefly, from May 2022 to April 2023, wastewater samples were collected biweekly from eight sampling locations across six wastewater treatment plants (WWTPs). The corresponding sewersheds are delineated in **Figure 1A**. Facility 4 is a 24-hr composite sample for the combined sewershed spanning Facility 4A and Facility 4B, which were also independently monitored using grab samples collected from the respective sewer trunk lines prior to their entry into Facility 4.

These samples were collected directly into amber glass vials containing 50 mg/L of ascorbic acid (oxidant quenching, albeit not needed for this study) and 1 g/L of sodium azide (biological preservation). Samples were briefly stored at 4°C prior to processing and analysis, typically within 1-2 days, and the target analytes included 17 PPCPs and 22 HRS, including major metabolites. Sample processing and analysis for PPCPs included automated solid phase extraction (ASPE) and injection of methanol extracts, while HRS analysis involved direct injection of 10-fold diluted aqueous samples. PPCP analysis was conducted using a SCIEX API 4000-series mass spectrometer (Redwood City, CA), employing both negative and positive electrospray ionization (ESI) in multiple reaction monitoring (MRM) mode. Drug analytes were tested on a SCIEX 6500 QTRAP mass spectrometer (Redwood City, CA, USA), focusing on positive ESI in MRM mode.

Five analytes, including delta-9-tetrahydrocannabinol (THC), heroin, 3,4-methylenedioxyamphetamine (MDA), norcocaine, and triclocarban, were detected in fewer than 10% of the samples in this study, so our analysis focused on the remaining 34 analytes (16 PPCPs and 18 HRS). This was in part due to aqueous instability (e.g., THC and heroin) and/or insufficient sensitivity (e.g., MDA and norcocaine) for certain analytes. However, the target compound list included other relevant analytes that could inform consumption patterns for the parent compounds of interest. For example, the major metabolites THC-COOH and THC-OH were used to assess THC consumption; the major metabolite 6-acetylmorphine was used to assess heroin use; and cocaine, benzoylecgonine, ecgonine methyl ester, and ecgonine served as alternatives to norcocaine. 3,4-Methylenedioxymethamphetamine (MDMA) is sufficiently stable as a parent compound to assess consumption directly (i.e., rather than using MDA). Finally, triclocarban was banned by the U.S. Food and Drug Administration (FDA) in September 2016^31^.

### Unsupervised clustering

To evaluate PPCP and HRS usage patterns across various facilities, we conducted an unsupervised clustering analysis on all 34 analytes, assessed at 26 distinct time points as features for each facility. We developed a similarity matrix for the facilities by calculating pairwise Pearson’s correlations, followed by hierarchical clustering to determine the similarities and differences among them. For an unbiased evaluation of the interconnections between different usage patterns, we carried out a separate hierarchical clustering analysis on the analytes, considering their concentrations at 26 time points across all six facilities (eight sampling locations). This approach helped establish similarities among analytes based on Pearson’s correlation measures. We applied this methodology for both PPCPs and HRS, enabling a comprehensive analysis of their respective usage patterns.

### Statistical analyses: Linear mixed effect model

To assess temporal changes and differences across facilities for each analyte, we employed a linear mixed effect (LME) model. This model incorporated fixed effects for the location (eight sampling locations), time (26 time points), and their interaction (location x time). Random effects included the intercept and time variation by location. To account for multiple comparisons (N_drug_x3), we applied a false discovery rate (FDR) correction method (*p_FDR_*) to the raw *p*-values for both main and interaction effects. To explore whether usage patterns vary based on population background and neighborhood context, our focus was on analytes showing significant location effects or interaction effects in the LME model. Post-hoc two-sample t-tests between Facilities 2 vs. 5 and Facilities 2 vs. 6 were conducted to analyze differences in usage patterns in relation to Area Deprivation Index (ADI) and Rural-Urban Commuting Area (RUCA) code variations, respectively. For all sampling locations, another LME model was utilized to examine the relationship between usage patterns and ADI scores, calculated as the average across all zip codes each facility covers. Here, we used individual analyte concentrations at each time point, rather than temporal averages, to enhance statistical power. The fixed effects in this second model were ADI and time, while the random effects remained consistent with the first model. All statistical analyses were conducted in MATLAB 2022b (https://www.mathworks.com/).

## Figure Legends

**Supplementary Table 1.** Direct comparison of PPCP and HRS loading (mg/day-person) at Facility 1 during the same months in 2010 and 2023.

| Drug | PPCP/HRS | Category | 2/7/2010 | 3/7/2010 | 2/6/2023 | 3/6/2023 | Increase in usage |
| --- | --- | --- | --- | --- | --- | --- | --- |
| Acetaminophen | PPCP | OTC Pain Reliever |  |  | 86.82 | 91.16 |  |
| Ibuprofen | PPCP | NSAID |  |  | 11.72 | 13.46 |  |
| Naproxen | PPCP | NSAID |  |  | 7.81 | 8.68 |  |
| Atenolol | PPCP | Licit Drug | 0.79 | 0.78 | 0.61 | 0.78 | -11.87% |
| Carbamazepine | PPCP | Licit Drug | 0.04 | 0.04 | 0.07 | 0.06 | 48.25% |
| Fluoxetine | PPCP | Licit Drug |  |  | 0.03 | 0.03 |  |
| Gemfibrozil | PPCP | Licit Drug |  |  | 0.61 | 0.61 |  |
| Meprobamate | PPCP | Licit Drug | 0.35 | 0.34 | 0.06 | 0.08 | -80.51% |
| Primidone | PPCP | Licit Drug | 0.08 | 0.05 | 0.10 | 0.10 | 59.58% |
| Sulfamethoxazole | PPCP | Licit Drug | 0.40 | 0.46 | 0.41 | 0.48 | 3.19% |
| Trimethoprim | PPCP | Licit Drug | 0.28 | 0.29 | 0.23 | 0.23 | -21.25% |
| Caffeine | PPCP | Daily |  |  | 47.75 | 56.43 |  |
| DEET | PPCP | Daily | 0.07 | 0.08 | 0.08 | 0.20 | 78.17% |
| Sucralose | PPCP | Daily |  |  | 43.41 | 56.43 |  |
| TCEP | PPCP | Daily | 0.13 | 0.17 |  |  |  |
| Triclosan | PPCP | Daily |  |  |  |  |  |
| Acetylmorphine | HRS | Opioid |  |  | 0.02 | 0.02 |  |
| Codeine | HRS | Opioid |  |  | 0.07 | 0.07 |  |
| EDDP | HRS | Opioid |  |  | 0.07 | 0.07 |  |
| Hydrocodone | HRS | Opioid |  |  | 0.06 | 0.08 |  |
| MDA | HRS | Opioid | 0.02 | 0.02 |  |  |  |
| MDMA | HRS | Opioid | 0.12 | 0.11 | 0.05 | 0.07 | -46.60% |
| Methadone | HRS | Opioid |  |  |  | 0.02 |  |
| Morphine | HRS | Opioid | 0.26 | 0.30 | 0.32 | 0.25 | 2.08% |
| Norcocaine | HRS | Opioid | 0.01 | 0.01 |  |  |  |
| Norfentanyl | HRS | Opioid |  |  | 0.04 | 0.04 |  |
| Oxycodone | HRS | Opioid |  |  | 0.06 | 0.06 |  |
| Tramadol | HRS | Opioid |  |  | 0.16 | 0.17 |  |
| Amphetamine | HRS | Stimulant | 0.13 | 0.14 | 0.30 | 0.31 | 128.17% |
| Benzoyllecgonine | HRS | Stimulant | 0.81 | 0.55 | 1.13 | 1.48 | 90.78% |
| Cocaine | HRS | Stimulant | 0.34 | 0.33 | 0.48 | 0.65 | 69.11% |
| Ecgonine | HRS | Stimulant | 0.31 | 0.30 | 0.13 | 0.17 | -48.89% |
| Ecgonine methyl ester | HRS | Stimulant | 0.18 | 0.15 | 0.40 | 0.52 | 175.47% |
| Methamphetamine | HRS | Stimulant | 0.92 | 1.04 | 2.69 | 2.78 | 180.21% |
| THC-COOH | HRS | Marijuana |  |  | 1.95 | 2.04 |  |
| THC-OH | HRS | Marijuana |  |  | 0.65 | 0.78 |  |

**Supplementary Figure 1.**
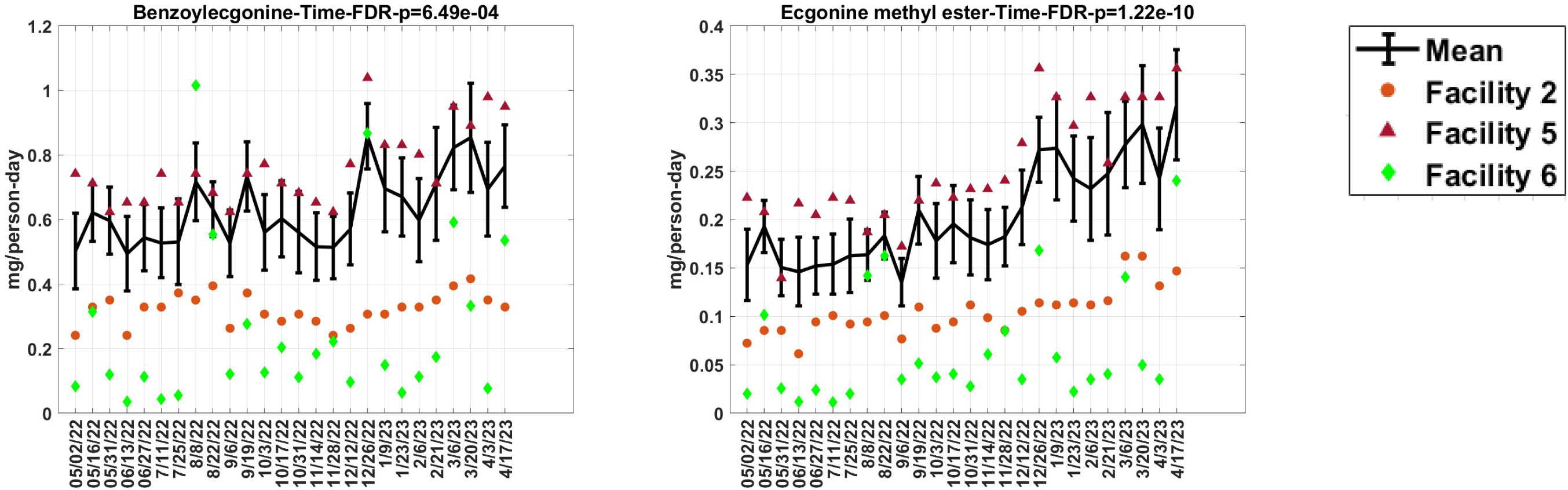
Metabolites of cocaine such as benzoylecgonine and ecgonine methyl ester increase (mg/person-day) over time from 2022-2023.

**Supplementary Figure 2.**
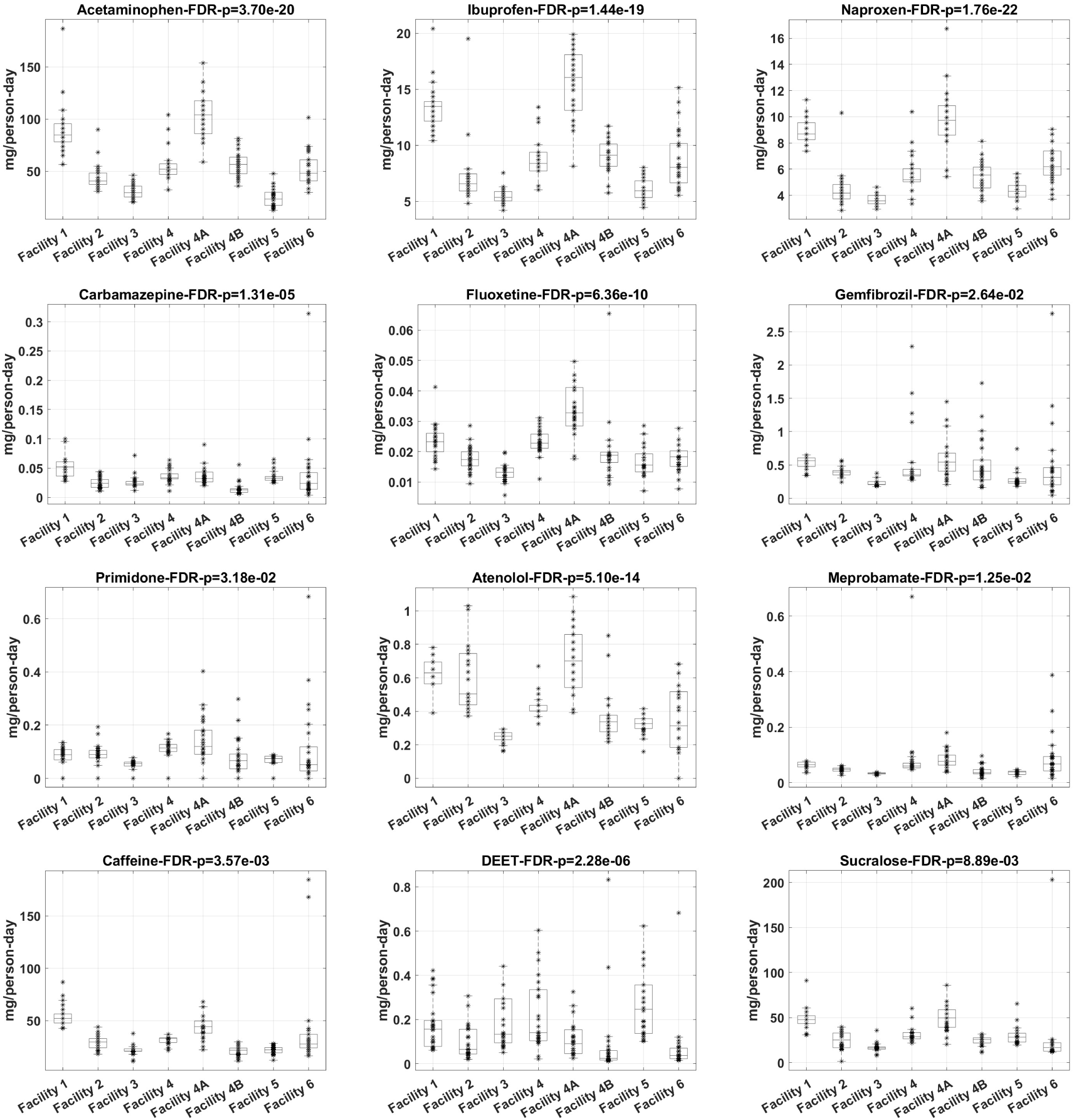
Usage patterns of 12 PPCPs, with a significant location effect in the linear mixed effect model, in wastewater from the eight sampling locations across six WWTP facilities.

**Supplementary Figure 3.**
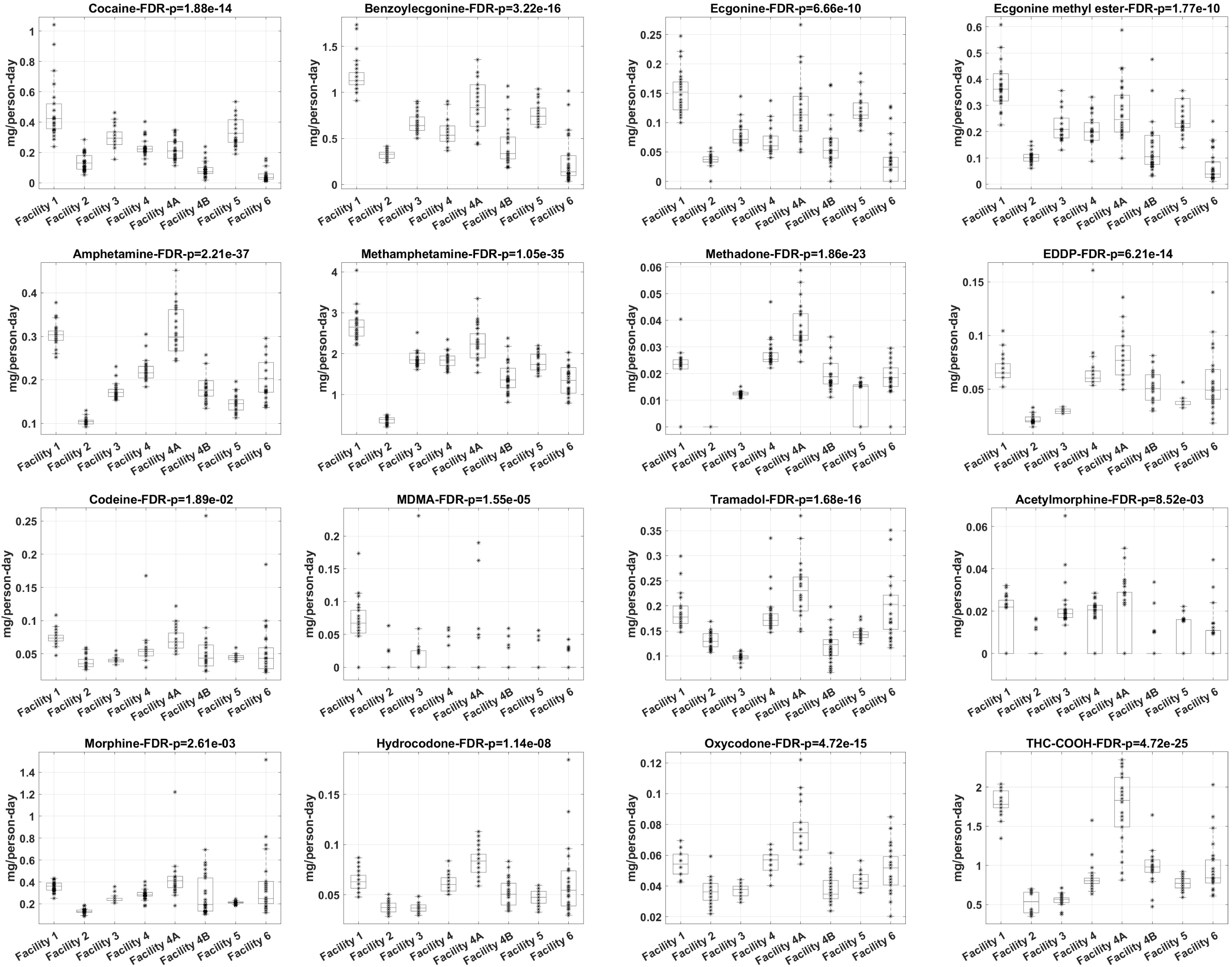
Usage patterns of 16 HRS analytes, with a significant location effect in the linear mixed effect model, in wastewater from the eight sampling locations across six WWTP facilities.

**Supplementary Figure 4.**
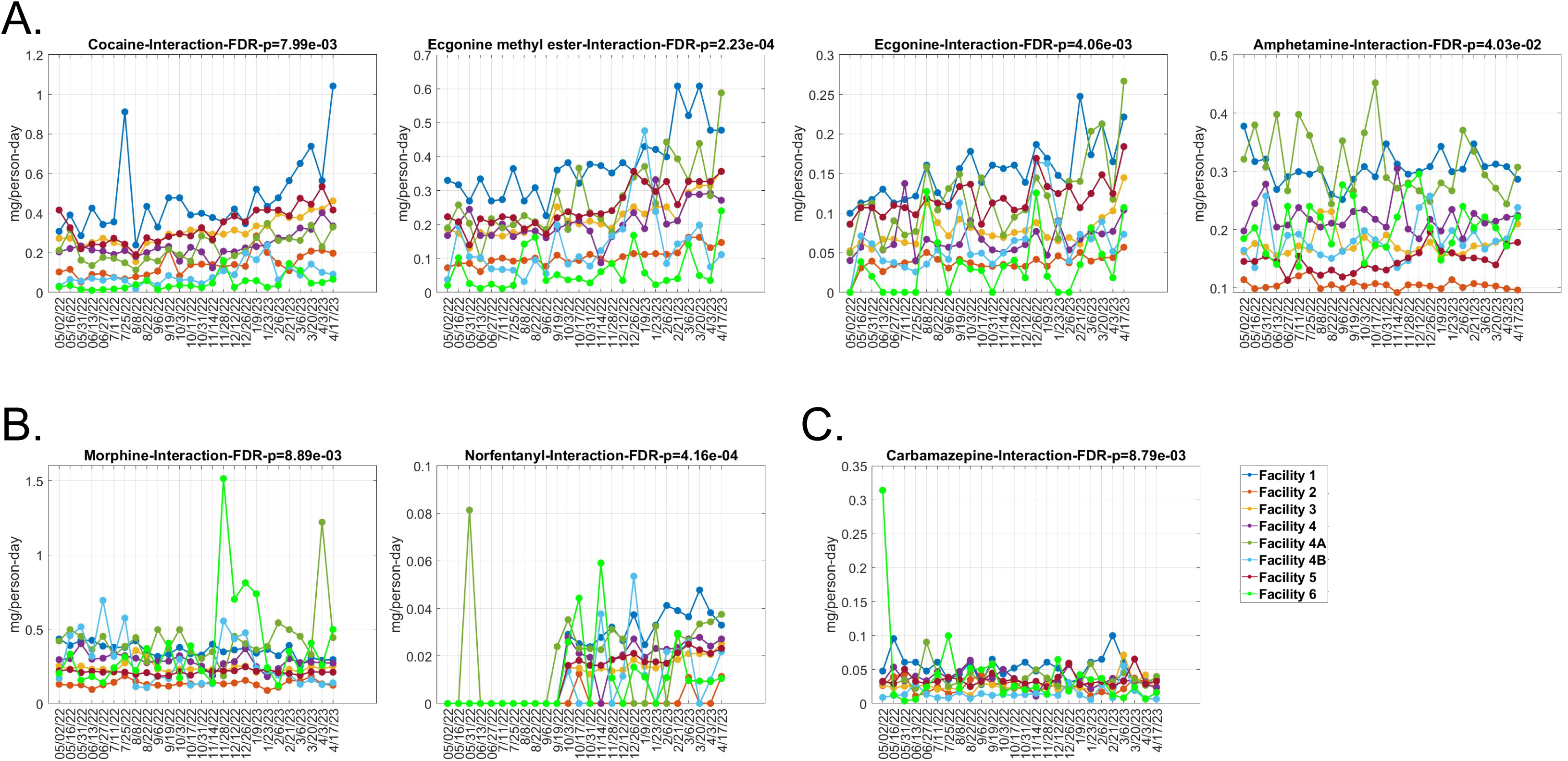
Temporal usage patterns across eight sampling locations of seven analytes with a significant location x time effect in the linear mixed effect model, including **(A)** four stimulants, **(B)** two opioids, and **(C)** one PPCP.

## Data Availability

All data produced in the present study are available upon reasonable request to the authors

## Acknowledgments

VV, ECO are supported by NIH grants: GM103440 and MH109706 and a CARES Act grant from the Nevada Governor’s Office of Economic Development. VV, CL, DG, HK, and ECO are supported by Grant Number NH75OT000057-01-00 from the Centers for Disease Control and Prevention. XZ and DC are supported by the NIH grant RF1-AG071566. The project contents are solely the responsibility of the authors and do not necessarily represent the official views of the Centers for Disease Control and Prevention. We would like to acknowledge personnel at collaborating wastewater agencies for their assistance with samples.

